# Blood-based biomarkers across the Alzheimer’s disease continuum in older Korean population

**DOI:** 10.64898/2026.10.27.26366659

**Authors:** Sarang Kang, Yu Yong Choi, Minji Kim, Kyu Yeong Choi, Eun Hyun Seo, Hyundoo Hwang, Ji Yeon Chung, Jahae Kim, Ho-Chun Song, Hoowon Kim, Youngshik Choe, Gyungah R. Jun, Kun Ho Lee

## Abstract

**Background:** Plasma phosphorylated tau 217 (pTau217) is a robust biomarker of Alzheimer’s disease (AD) pathology. However, whether a single biomarker can adequately capture both clinical diagnosis and disease progression remains unclear. We investigated the complementary roles of plasma AD biomarkers and routinely measured metabolic markers across different stages of AD.

**Methods:** Using the Gwangju Alzheimer’s & Related Dementia (GARD) cohort, we analyzed cross-sectional and longitudinal data, including amyloid-PET, MRI, neuropsychological examinations, and *APOE* genotypes. Plasma pTau217, GFAP, and NfL levels were measured alongside routine biochemical markers to evaluate their associations with pathology and neurodegeneration.

**Results:** Plasma pTau217 best identified amyloid-PET positivity (AUC > 0.91). For AD diagnosis, adding cholesterol and glucose to pTau217 improved the AUC from 0.781 to 0.873. To predict progression in amyloid-positive individuals, glucose alone outperformed pTau217 (AUC 0.840 vs. 0.747), whereas adding GFAP yielded the strongest prognostic performance (AUC 0.867).

**Discussion:** Plasma pTau217 primarily reflects amyloid pathology, whereas metabolic markers provide complementary information related to the clinical expression and progression of the disease. Integrating biomarkers across distinct biological domains may improve risk stratification and prognostic assessment in AD.

## INTRODUCTION

Alzheimer’s disease (AD) is a progressive neurodegenerative disorder that poses a significant global health challenge. The quest for accurate and minimally invasive diagnostic tools has led to an increasing interest in blood or plasma-based biomarkers, which offer the potential for early detection and monitoring of AD pathology before the onset of clinical symptoms.(1) Promising plasma biomarkers include phosphorylated tau 217 (pTau217), glial fibrillary acidic protein (GFAP), and neurofilament light chain (NfL).(2–5) Specifically, plasma pTau217 reflects the core pathology of amyloid and tau protein accumulation, hallmarks of AD pathology, with potentially higher diagnostic accuracy than other plasma biomarkers. However, the link between AD pathology and clinical symptoms varies, with many amyloid-positive individuals remaining cognitively unimpaired, while others quickly develop dementia.

Recent studies suggest that biomarkers reflecting distinct biological processes, including astrocytic activation, neurodegeneration, and metabolic dysfunction, may provide information beyond core AD pathology.(6–8) Plasma GFAP has been associated with cognitive decline and incident dementia, while metabolic abnormalities such as dyslipidemia and hyperglycemia have been linked to accelerated disease progression.(8, 9) However, whether these biomarkers provide complementary diagnostic and prognostic information beyond pTau217 remains unclear.

This study investigated the predictive value of plasma pTau217, GFAP, and NfL levels for amyloid positivity in a well-characterized longitudinal Korean cohort from the Gwangju Alzheimer’s and Related Dementias (GARD) study.(10) To enhance the sensitivity for detecting the clinical diagnosis and progression of AD, we also evaluated metabolic markers, including plasma cholesterol and glucose levels, in combination with these biomarkers. We hypothesized that distinct biomarker domains differentially contribute to amyloid pathology, clinical diagnosis, and disease progression.

## METHODS

### Participants

The participants were enrolled from the Gwangju Alzheimer’s and Related Dementias (GARD) research center, a large-scale dementia cohort in South Korea.(10) Individuals aged ≥60 years were recruited from the local community and underwent standardized annual follow-up evaluations, including neuropsychological assessment, brain imaging, and blood sampling. Detailed information on the inclusion and exclusion criteria, follow-up assessments, and available multimodal data has been previously described.(10) In this study, age- and sex- matched participants were selected from the GARD cohort based on the availability of comprehensive neuropsychological assessments, neuroimaging, and blood samples. Participants were included only if complete clinical and biomarker data were available, ensuring diagnostic confirmation and consistency in the model development.

### Phenotype Assessment

All participants underwent neurological examinations, including the Korean Mini-Mental State Examination (K-MMSE) and the Seoul Neuropsychological Screening Battery (SNSB).(11) Additionally, all participants underwent pathological examinations, including 3T magnetic resonance imaging (MRI; Skyra, Siemens Healthineers Ltd., Seoul, Republic of Korea) and ^18^F-Florbetaben (FBB) beta-amyloid positron emission tomography (PET) scans. Brain MRI scans of the GARD cohort participants were obtained using a 3.0 T Skyra (Siemens) scanner with a 20-channel head coil in the MPRAGE sagittal orientation.

Comprehensive diagnostic procedures have been previously described in detail.(10) Briefly, clinical diagnoses were established by integrating cognitive test performance, functional ability, and medical history with structural neuroimaging findings. Diagnostic classification followed established guidelines, including the Clinical Dementia Rating (CDR), the Diagnostic and Statistical Manual of mental disorders, Fifth Edition (DSM-5) criteria, and the National Institute on Aging and the Alzheimer’s Association (NIA-AA) research framework.(10) Structural MRI was incorporated into the diagnostic workup to identify vascular pathology and other structural abnormalities. To reduce the likelihood of including vascular dementia, individuals with a white matter hyperintensity score of ≥2 or with imaging evidence of infarction or hemorrhage were excluded from the study. Participants were subsequently assigned to one of three diagnostic categories: cognitively unimpaired (CU), mild cognitive impairment (MCI), or Alzheimer’s dementia. Brain amyloid burden was assessed using FBB-PET, and β-amyloid deposition was quantified using standardized uptake value ratios (SUVr). Amyloid positivity was determined by a board-certified nuclear medicine physician based on a visual assessment of PET images in conjunction with SUVr values, and participants were subsequently classified as amyloid-negative or amyloid-positive.

We utilized longitudinal data of up to 10 years from the GARD cohort to evaluate changes in clinical diagnosis over time. To ensure prognostic relevance in the context of AD pathology, longitudinal analyses were restricted to amyloid-positive individuals. We defined progression as a retrospective stage transition at the last examination, from CU to AD or from MCI to AD, between the baseline and the most recent visit concurrent with biomarker measurement. Participants who met these criteria were classified as converters (status=1), whereas those who remained clinically stable were classified as non-converters (status=0). Patients with dementia at baseline were excluded from the conversion analyses. All diagnoses at both baseline and follow-up examinations were determined by consensus among board-certified neurologists and psychiatrists based on comprehensive clinical evaluations.

### Plasma and blood biomarker assessment

Fasting blood samples were collected in plastic collection tubes containing ethylenediamine tetra-acetic acid (EDTA) at the last examination. The samples were centrifuged twice at 2,000 g and 3,000 g at 4 and only the supernatant was extracted. Plasma was separated and stored in a −80 degrees deep freezer. Plasma biomarkers were measured using a single-molecule array (Simoa) platform (Quanterix, Billerica, MA, USA)(12). All measurements were performed at Bredis Co., Ltd., using commercially available assay kits. Plasma p-tau217 concentrations were quantified using the Simoa ALZpath pTau217 Advantage PLUS assay kit (104570; Quanterix). Plasma GFAP and NfL levels were measured using the Simoa Neurology 2-plex B assay kit (103520; Quanterix). The measurements were performed separately in three experimental rounds. The intra-assay coefficients of variation were < 10%.

Blood biochemical markers were selected based on the standard Korean health screening items and the GARD cohort protocol, including thrombocytes, hemoglobin, hematocrit, total cholesterol, low-density lipoprotein cholesterol (LDL), high-density lipoprotein cholesterol (HDL), triglyceride, aspartate aminotransferase (AST), alanine aminotransferase (ALT), blood urea nitrogen (BUN), creatinine, glycated hemoglobin (HbA1C), thyroid stimulation hormone (TSH), and glucose.

### Statistical Analysis

Before analysis, biomarker values exceeding three standard deviations (SD) from the mean were excluded as extreme outliers. This resulted in the removal of 4 values for pTau217, 5 for GFAP, and 20 for NfL. The baseline characteristics of the remaining cohort are summarized as means ± standard deviations (SDs) for continuous variables and as counts (n) with percentages (%) for categorical variables. We then assessed the differences between the CU, MCI, and AD groups using Analysis of Covariance (ANCOVA). These models were adjusted for age, sex, and education when comparing neuropsychological tests, and for age, sex, and intracranial volume (ICV) when comparing neuroimaging phenotypes.

We conducted associations of AD-related biomarkers including plasma levels of pTau217, GFAP, and NfL for AD diagnosis and amyloid positivity as binary outcomes using logistic regression models and for neuropsychological scores and neuroimaging phenotypes as quantitative outcomes using linear regression models. These models were adjusted for age, sex, and education for binary outcomes and quantitative neuropsychological scores, and for age, sex, and intracranial volume (ICV) for quantitative neuroimaging phenotypes. We further evaluated the predictive performance of the biomarkers for amyloid positivity and AD diagnosis using receiver operating characteristic (ROC) curve analysis, adjusting for age, sex, and education. We calculated the area under the curve (AUC) to predict amyloid positivity and AD diagnosis in all participants and within the CU or MCI subgroups to assess the early detection of AD pathology. To evaluate the applicability of a previously established plasma pTau217 cut-off in the present study, the threshold of 0.42 pg/mL reported by Ashton et al.(13), which was derived using the same ALZpath Simoa assay, was externally validated against amyloid positivity. The diagnostic performance of the threshold was evaluated in terms of sensitivity, specificity, positive predictive value (PPV), and negative predictive value (NPV). To further assess the optimal threshold within the present cohort, an additional ROC-based optimization was performed, and the best cutoff was determined using the Youden index.

To identify additional predictive blood markers in the amyloid-positive group, we conducted a secondary analysis excluding amyloid-negative individuals. We selected promising blood biochemical markers by performing ANCOVA on group comparisons between CU, MCI, and AD participants, adjusting for age, sex, and level of education. Subsequently, significant markers were added to the primary AD-related biomarkers to test the associations between the combined blood markers and AD diagnosis (CU vs. AD), which were further adjusted for medication status for diabetes or hyperlipidemia treatments. Bonferroni correction at p<0.005 was applied to account for multiple test comparisons.

## RESULTS

### Phenotypic characteristics for AD status and amyloid positivity

The participant selection process is illustrated in **Supplementary Figure 1**. A total of 725 Korean participants were included in the analysis, consisting of 442 CU, 224 MCI, and 59 AD subjects (**Supplementary Table 1**). The diagnosis groups were matched for age, sex, and *APOE* ε4 status between groups (p>0.05). However, years of education were nominally significant across groups (p=0.013) and were included as a covariate in subsequent analyses. We observed that twice as much as the number of non-AD subjects took medications related to hyperlipidemia or diabetes (p=0.004) than AD cases (**Supplementary Table 1**). AD-related endophenotypes, including neuropsychological battery tests and neuroimaging scans, were significantly different between the AD diagnosis groups (**Supplementary Tables 1 and 2**). Among these endophenotypes, only cognitive tests in the memory and executive function domains were significantly associated with amyloid positivity after multiple testing correction at p<0.005 (**Supplementary Table 2**).

### Blood-based biomarkers for amyloid positivity and AD diagnosis using all subjects

We observed significant differences in the levels of all three plasma biomarkers between the CU and AD groups (**Figure 1A and Supplementary Table 1**). Among these, pTau217 was the only biomarker that showed significant differences across all three comparisons (CU vs. MCI, MCI vs. AD, and CU vs. AD) (**Figure 1A**). Both pTau217 and GFAP significantly differentiated amyloid-positive and amyloid-negative individuals in all three AD diagnostic groups (**Figure 1B and Supplementary Table 2**), indicating their strong discriminatory ability for amyloid pathology. All three biomarkers were significantly associated with cognitive tests, hippocampal volume, and entorhinal thickness, even after multiple testing corrections in amyloid-positive participants (**Supplementary Tables 3-5**).

**Figure 1.**
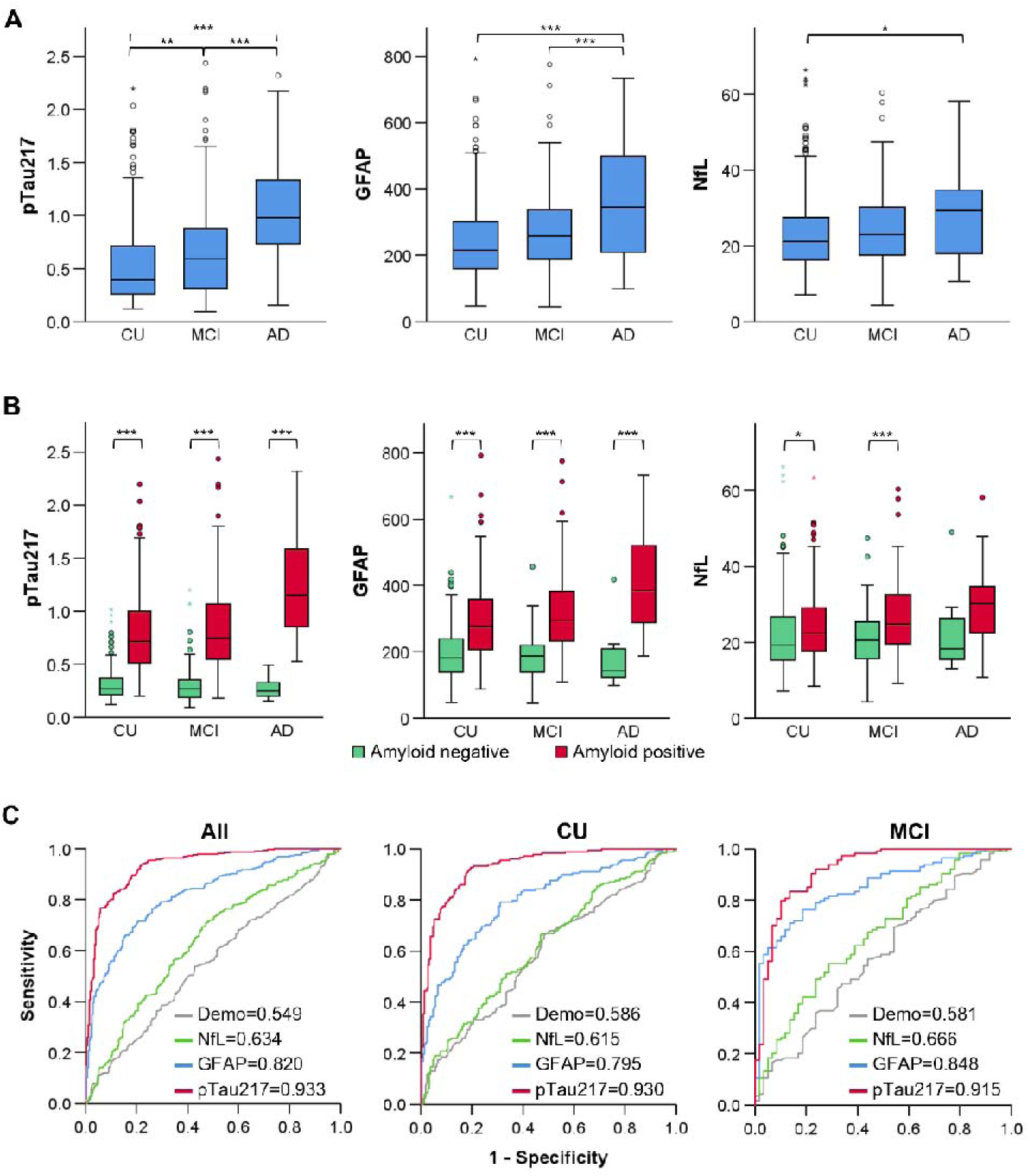
Distribution and predictive performance of plasma biomarkers across diagnostic and amyloid status. **(A)** Levels of pTau217, GFAP, and NfL by AD status. **(B)** Distribution of plasma biomarkers stratified by amyloid positivity within each AD status. **(C)** Receiver operating characteristic (ROC) curves for predicting amyloid positivity using plasma biomarkers, adjusted for demographics (Demo) including age, sex, education level. Asterisks indicate statistical significance: \**p*<0.05, \*\**p*<0.01, \*\*\**p*<0.001.

The predictive performance of the AD-related biomarkers was assessed for amyloid positivity (**Figure 1C**) and clinical diagnosis groups (**Supplementary Table 6**) adjusted for age, sex, and education (demographics) in the total sample. When evaluating biomarkers for predicting amyloid positivity, plasma pTau217 demonstrated the best predictive accuracy, yielding an AUC of 0.933 (**Figure 1C**). This was substantially higher than the performance of GFAP (AUC=0.820) and NfL (AUC=0.634) in all subjects. This high performance for pTau217 remained consistent even in preclinical stages, including both CU (AUC = 0.930) and MCI (AUC = 0.915).

To further validate the clinical utility of plasma pTau217 in this cohort, we applied the previously established cut-off of 0.42 pg/mL, which yielded a sensitivity of 86.4%, specificity of 85.7%, PPV of 88.4%, and NPV of 83.4%. ROC-based optimization within the present cohort identified a slightly lower threshold of 0.395 pg/mL (sensitivity 88.9%, specificity 82.6%, PPV 86.5%, NPV 85.6%), which did not substantially differ in overall diagnostic performance from the published threshold. These findings suggest that the pre-established cutoff generalizes well to the Korean population.

We next assessed the predictive performance of AD-related biomarkers in distinguishing between different clinical stages. pTau217 was the top-performing biomarker for discriminating CU from AD (AUC=0.790), and adding other biomarkers to the model did not further improve this diagnostic performance (**Supplementary Table 6**). When discriminating MCI from AD, the predictive performance of pTau217 was reduced (AUC=0.713), but adding NfL to the model slightly improved the performance (AUC=0.723). No biomarker was effective at the earliest preclinical stage to distinguish between the CU and MCI stages (AUC<0.61) (**Supplementary Table 6**).

None of the routine biochemical markers were significantly associated with amyloid positivity (**Supplementary Table 7**). We observed a significant association between cholesterol, LDL, and glucose levels in CU and AD after multiple testing corrections (**Supplementary Table 7**). The predictive performance of a model containing cholesterol and glucose after adjusting for age, sex, education, and medication use was modest between CU and AD (AUC=0.765) and between MCI and AD (AUC=0.712) (**Supplementary Table 8**). However, the AUC between CU and MCI was low, suggesting that these markers may be more closely linked to the onset of clinical symptoms than to early amyloid accumulation.

### Combined blood-based biomarkers for AD diagnosis in amyloid-positive individuals

Our analysis of CU individuals revealed a distinct pattern of biomarker changes corresponding to early amyloid pathology (**Table 1 and Figure 2**). Specifically, both plasma pTau217 and GFAP levels were already elevated in amyloid-positive (CU+) individuals compared to amyloid-negative (CU-) individuals (**Table 1 and Figure 2A**). Plasma pTau217 showed dramatical early increase in CU+ (mean=0.80) compared to CU-(mean=0.30) individuals. In contrast, neither plasma NfL nor any of the routine biochemical markers showed a significant difference between the CU+ and CU-groups, suggesting that they are not involved in the initial phase of amyloid accumulation and likely change only at later stages of the disease (**Table 1 and Figure 2**).

**Figure 2.**
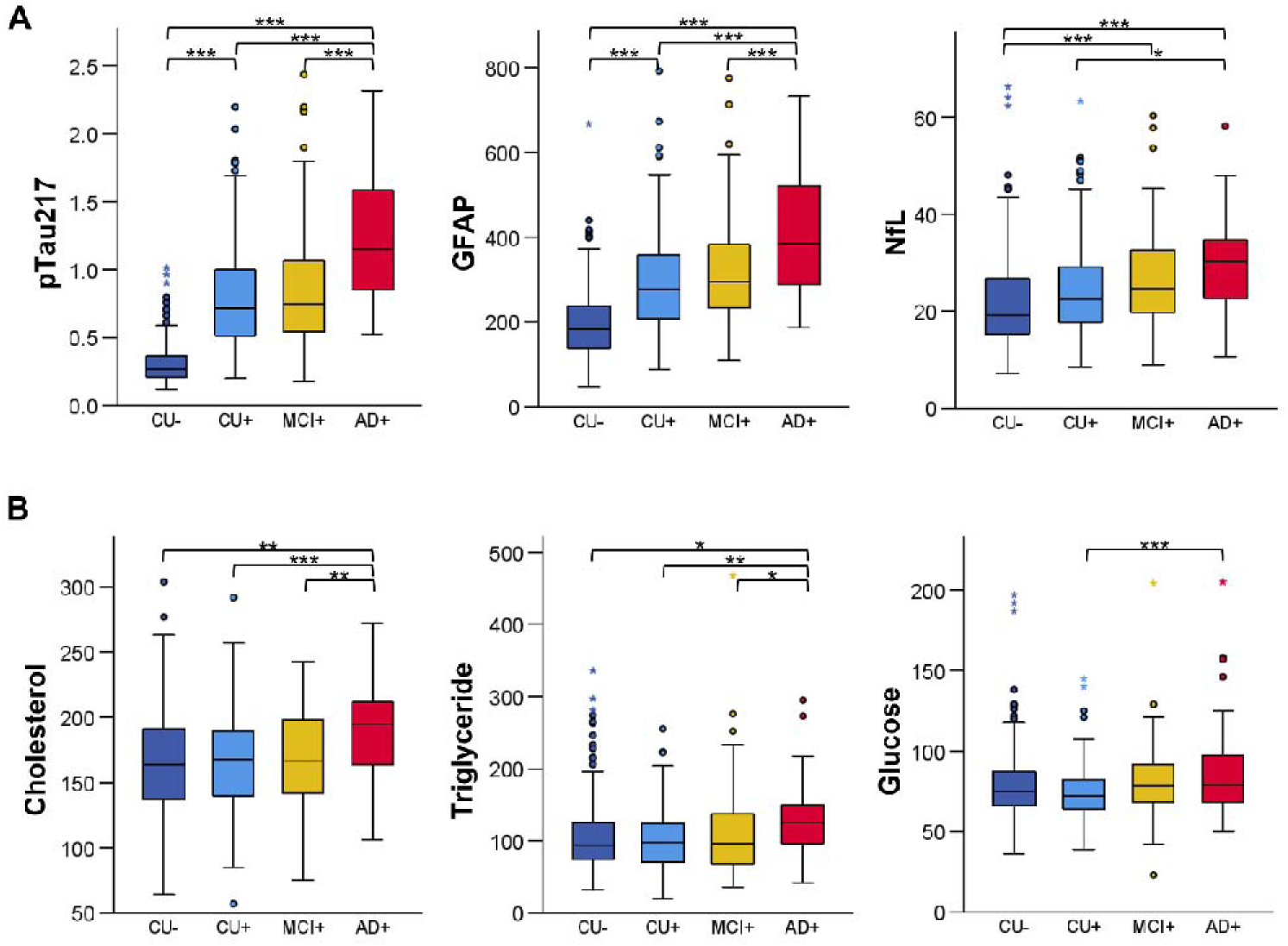
Distribution of plasma biomarkers and blood biochemical markers across amyloid-diagnostic groups. **(A)** Boxplots of plasma biomarkers across four group defined by cognitive status and amyloid positivity: CU amyloid-negative (CU-), CU amyloid-positive (CU+), MCI amyloid-positive (MCI+), and AD amyloid-positive (AD+). **(B)** Distribution of blood biochemical markers across the same diagnostic subgroups. Asterisks indicate statistical significance: *p<0.05, **p<0.01, ***p<0.001.

Further evaluation of these biomarkers restricted to the amyloid-positive groups (CU+, MCI+, and AD+) showed that all three AD biomarkers (pTau217, GFAP, and NfL), as well as several biochemical markers (total cholesterol, triglyceride, LDL, and glucose), were significantly elevated in individuals with dementia (AD+) compared to asymptomatic cognitively unimpaired (CU+) subjects (**Figures 2A and 2B**). Of these, pTau217, GFAP, total cholesterol, and triglycerides also increased significantly from MCI+ to the AD+ stages. In amyloid-positive individuals, the predictive performances of glucose and pTau217 varied according to the clinical stage. After adjusting for demographics and medication, glucose showed a slightly better AUC than pTau217 in both CU+ and AD+ individuals (0.783 vs. 0.777; **Figure 3A**) and CU+ and MCI+ individuals (0.575 vs. 0.661; **Figure 3B**) (**Supplementary Table 9**). Conversely, for discriminating between MCI+ and AD+, pTau217 was a better predictor (AUC: 0.737 vs. 0.696) (**Figure 3C and Supplementary Table 9**). The integrated model substantially improved performance, and combining glucose and pTau217 increased the AUC to 0.861 for the CU+ vs. AD+ comparison. The comprehensive model, which included additional cholesterol and GFAP, achieved the best performance, with an AUC of 0.869 (**Supplementary Table 9**).

**Figure 3.**
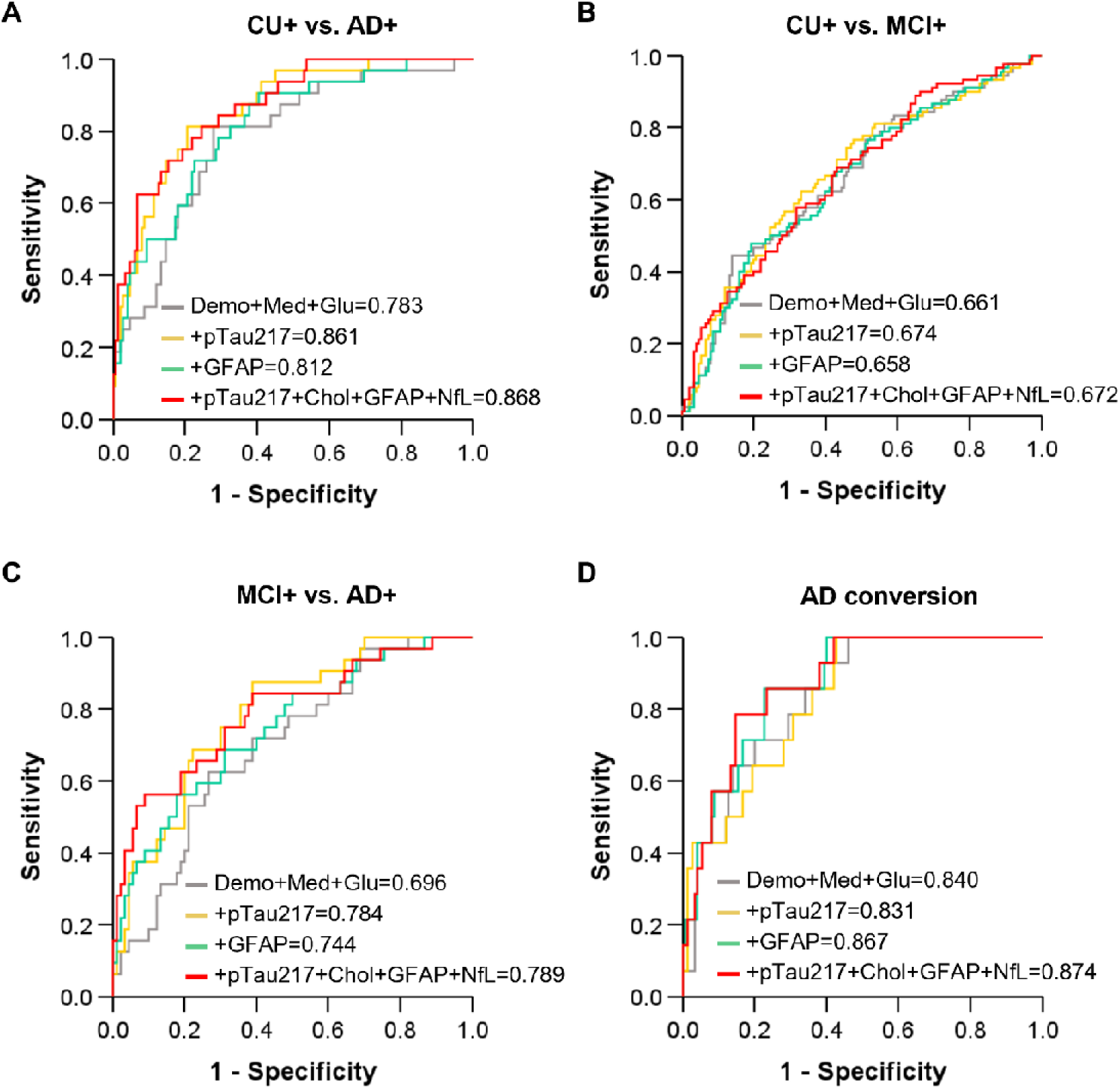
Predictive performance of novel blood biochemical markers for AD status, and AD conversion in amyloid positive subgroup. ROC curve for AD status and conversion using plasma biomarkers and blood biochemical markers, adjusted for demographics (age, sex, and education level) and medication. Variables are abbreviated as follows: Demo, demographics; Medi, medication; Glu, glucose; Chol, cholesterol.

### Combined blood-based biomarkers for AD progression in amyloid-positive individuals

During a 4–5-year follow-up period, 4 CU+ and 14 MCI+ individuals converted to AD, whereas no CU- and only one MCI-individuals progressed to AD (**Supplementary Table 10**). When comparing conversion rates among amyloid-positive individuals from CU+ or MCI+ to AD+, glucose outperformed pTau217 after adjusting for demographics and medication (AUC: 0.840 vs. 0.747) (**Supplementary Table 11**). Adding GFAP to the glucose model further improved performance to an AUC of 0.867, which was better from the model adding pTau217 (AUC: 0.867 vs. 0.831) (**Figure 3D**). The comprehensive model containing all five markers showed the best performance achieving AUC of 0.874 (**Supplementary Table 11**).

## DISCUSSION

Plasma pTau217 has emerged as one of the most accurate blood-based biomarkers for AD, primarily reflecting cerebral amyloid pathology. (7, 13–15) In the present study, we extend these observations by demonstrating that different biomarker domains provide complementary information across distinct stages of AD. While pTau217 showed the strongest performance for identifying amyloid-positive individuals, metabolic markers contributed additional information for clinical diagnosis and disease progression. These findings suggest that no single biomarker adequately captures the full spectrum of AD biology and support a multi-domain biomarker framework for risk stratification and prognosis.

The discrepancy between AD pathology and clinical manifestation is increasingly recognized as a major challenge in the field. Although amyloid accumulation can begin decades before symptom onset, substantial heterogeneity exists in the timing and rate of progression to dementia. Consistent with this concept, pTau217 was highly informative for amyloid pathology in our cohort, whereas the addition of total cholesterol and glucose significantly improved discrimination between amyloid-positive cognitively unimpaired individuals and patients with AD dementia. These findings suggest that metabolic dysfunction may contribute information related to the clinical expression of AD pathology beyond amyloid burden alone.

The inclusion of total cholesterol and glucose, together with pTau217, revealed a critical intersection between metabolic risk factors and neurodegeneration. A high level of cholesterol in the blood, known as hyperlipidemia, can cause plaque within the blood vessels, leading to atherosclerosis.(16) This condition can lead to cardiovascular disease, such as angina, myocardial infarction, stroke, and AD.(17, 18) A significant number of studies have focused on the intricate relationship between hyperlipidemia, genes related to cholesterol metabolism (such as the APOE gene), and the pathogenesis of AD.(19, 20) Elevated fasting glucose levels are a hallmark of diabetes, a condition that can lead to complications including cardiovascular disease, nephropathy, and neuropathy.(21–23) In addition, accumulating evidence has indicated that insulin resistance and glucose metabolism disorders in the brain play a role in the pathological mechanisms underlying AD.(24–26) The mechanisms are thought to be multifactorial, involving contributions to cerebrovascular pathology, inflammation, and direct effects on amyloid precursor protein (APP) processing and amyloid-beta clearance.(27) Our findings support the concept that metabolic factors contribute complementary information beyond AD-specific biomarkers when evaluating disease severity and clinical status.

Among amyloid-positive individuals, GFAP demonstrated greater utility for predicting clinical progression than pTau217. Plasma GFAP is a sensitive marker of astrocyte reactivity associated with amyloid pathology, with increases already in the preclinical phase, thus supporting its role as a risk and progression indicator.(6, 28–30) Our findings further support its role as a prognostic biomarker, suggesting that astrocytic responses may be more closely linked to subsequent clinical deterioration than markers primarily reflecting amyloid burden.

The clinical implications of this study for predicting amyloid pathology in preclinical stages as well as AD progression are significant by facilitating prevention and early intervention in enriched individuals with fast decliners. Rather than relying on a single biomarker for all clinical purposes, different biomarker domains may be more informative depending on the intended application. pTau217 appears most useful for identifying underlying AD pathology, whereas metabolic markers may improve prediction of clinical diagnosis and future progression. The advantages of high accessibility and low invasiveness offer unique opportunities to screen a larger population of individuals at high risk for dementia, assess prognosis before the onset of brain damage, and develop personalized intervention strategies This study has several limitations. First, although the findings were statistically robust and the study was well-characterized, the cohort was drawn from a specific research setting, it may not fully represent the general population and limit the generalizability. Therefore, validation of our findings in more diverse, multi-ancestry, and population-based cohorts is an essential next step. Second, certain plasma markers, such as NfL, were selected based on post hoc associations, which may introduce potential selection bias. Third, the analyses were observational in nature and did not establish causal relationships between biochemical markers and AD-related outcomes. Fourth, while we included commonly used clinical covariates and adjusted for medication status, residual confounding due to unmeasured variables cannot be ruled out. Finally, although associations between peripheral biochemical markers and neurodegeneration were observed, the underlying biological mechanisms linking systemic metabolism to brain pathology remain to be elucidated.

Our study demonstrated that the progression of clinical symptoms in early stages of AD is a complex process influenced by metabolic risk factors in presence of the core pathologies of amyloid and tau. By combining pTau217 with readily available metabolic and neurodegenerative markers in blood, we significantly improved the prediction of both brain atrophy and clinical decline. This increased potential for clinical application of preclinical risk stratification, offering personalized and preventive strategies to managing AD.

## CONCLUSIONS

Plasma pTau217 was the strongest marker of cerebral amyloid pathology, whereas metabolic biomarkers provided complementary information for clinical diagnosis and disease progression. These findings support a multi-domain biomarker framework in which distinct biomarkers capture different aspects of AD biology rather than a single universal marker. Integrating AD-specific and metabolic biomarkers may improve risk stratification and prognostic assessment across the AD continuum.

## Data Availability

The data that support the findings of this study are available from GARD cohort but restrictions apply to the availability of these data, which were used under license for the current study, and so are not publicly available. Data are however available from the authors upon reasonable request and with permission of GARD cohort.

## List of abbreviations

AD: Alzheimer’s disease
AUC: Area under the curve
APOE: Apolipoprotein E
Aβ: Amyloid-beta
BUN: Blood urea nitrogen
CI: Confidence interval
CU: Cognitively unimpaired
GARD: Gwangju Alzheimer’s & Related Dementia
GFAP: Glial fibrillary acidic protein
HDL: High-density lipoprotein cholesterol
LDL: Low-density lipoprotein cholesterol
MCI: Mild cognitive impairment
MRI: Magnetic resonance imaging
NfL: Neurofilament light chain
NPV: Negative predictive value
PET: Positron emission tomography
PPV: Positive predictive value
pTau217: Phosphorylated tau 217
ROC: Receiver operating characteristic
SD: Standard deviation
Simoa: Single molecule array
TSH: Thyroid-stimulating hormone

## DECLARATIONS

## Ethics approval and consent to participate

This study was approved by the Institutional Review Boards of Chosun University Hospital (IRB No. CHOSUN 2019-10-022-025) and Chonnam National University Hospital (IRB No. CNUH-2019-279). All study participants, or their legal guardian, provided informed written consent prior to study enrollment.

## Consent for publication

Not applicable

## Competing interests

The authors declare that they have no competing interests

## Funding

This study was supported by the KBRI Basic Research Program through the Korea Brain Research Institute, funded by the Ministry of Science and ICT (26-BR-05-01), the “Korea National Institute of Health” research project (project No. 2025-ER1004-00), by the Technology Innovation Program (20022810, Development and Demonstration of a Digital System for the evaluation of geriatric Cognitive impairment) funded By the Ministry of Trade, Industry & Energy (MOTIE, Korea), by the Technology Innovation Program(or Industrial Strategic Technology Development Program)(RS-2024-00433283, Development of an AI-Driven Platform for Clinical Validation in Senior Healthcare Focused on Degenerative Brain Diseases) funded By the Ministry of Trade, Industry & Resources(MOTIR, Korea), by Brain Pool program funded by the Ministry of Science and ICT through the National Research Foundation of Korea (RS-2024-00407198), and by grant of the Korea Health Technology R&D Project through the Korea Health Industry Development Institute (KHIDI), funded by the Ministry of Health & Welfare, Republic of Korea (grant number: RS-2022-KH129598)..

## Authors’ contributions

KHL and GRJ designed and supervised the research. SK performed the data analyses, interpreted the data, and drafted the manuscript. YYC, MK, KYC, EHS, HH, JYC, JK, HCS, HK, and YC contributed to data acquisition and data curation. KHL, GRJ, and SK interpreted the data. GRJ and KHL critically revised the manuscript for important intellectual content. All authors have approved the submitted version.

## Acknowledgements

Not applicable

## Notes

### Competing Interest Statement

The authors have declared no competing interest.

