## Supplementary Figure and Tables for "Blood-based biomarkers across the Alzheimer’s disease continuum in older Korean population"

**Supplemental Online Content**

**Supplementary Figure 1.** Study design and analytical workflow

**Supplementary Table 1.** Baseline characteristics of phenotypes and plasma biomarkers in different diagnosis groups

**Supplementary Table 2.** Association with endophenotypes for AD status and Amyloid positivity as binary outcomes

**Supplementary Table 3.** Association for pTau217 and AD related endophenotypes in all, amyloid positive, and negative groups

**Supplementary Table 4.** Association for GFAP and AD related endophenotypes in all, amyloid positive, and negative groups

**Supplementary Table 5.** Association for NfL and AD related endophenotypes in all, amyloid-positive, and amyloid-negative subjects

**Supplementary Table 6.** Prediction of AD status using plasma biomarkers in all subjects

**Supplementary Table 7.** Association of blood biochemical markers for AD status and amyloid positivity in all subjects

**Supplementary Table 8.** Predictive performance in ROC curves of AD status using blood biochemical markers in all subjects

**Supplementary Table 9.** Predictive performance in ROC curves of AD status using pTau217 and blood biochemical markers in amyloid positive subgroup

**Supplementary Table 10.** Conversion to AD dementia during follow-up

**Supplementary Table 11.** AD conversion analysis using pTau217 and blood biochemical markers in amyloid positive subgroup

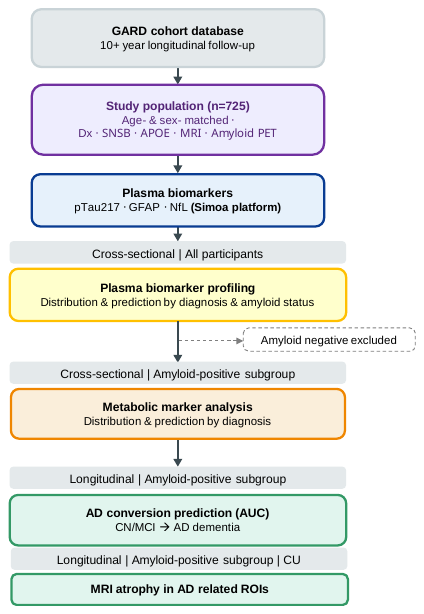

**Supplementary Figure 1. Study design and analytical workflow**. Flowchart illustrating participant selection and the analytical framework of the study. Participants were derived from the GARD longitudinal community-based dementia cohort with more than 12 years of follow-up. Age- and sex-matched individuals with available cognitive assessments, neuroimaging, and blood samples were included (n = 725). Plasma biomarkers (pTau217, GFAP, and NfL) were quantified using Simoa assays. Predictive modeling analyses were conducted using plasma biomarkers and blood biochemical markers. Longitudinal outcomes, including diagnostic conversion and brain atrophy progression, were retrieved from follow-up data to evaluate disease progression.

**Supplementary Table 1. Baseline characteristics of phenotypes and plasma biomarkers in different diagnosis groups**

| **Characteristics** | |  | **CU** | **MCI** | **AD** | **P** |
| --- | --- | --- | --- | --- | --- | --- |
| Total, N | |  | 442 | 224 | 59 |  |
| Age, mean (SD), y | | | 74.64 (6.2) | 74.09 (6.1) | 74.41 (7.7) | 0.573 |
| Sex, No. (%) | | |  |  |  |  |
|  | Female | | 230 (52.0) | 120 (53.6) | 32 (54.2) | 0.904 |
|  | Male | | 212 (48.0) | 104 (46.4) | 27 (55.8) |  |
| Education, mean (SD), y | | | 10.78 (4.5) | 10.69 (4.4) | 8.93 (4.6) | 0.013 |
| *APOE*, No. (%) | | |  |  |  |  |
|  | ε4 non-carrier | | 251 (56.8) | 126 (56.3) | 27 (45.8) | 0.193 |
|  | ε4 carrier | | 180 (40.7) | 97 (43.3) | 32 (54.2) |  |
| SUVr, mean (SD) | | | 1.34 (0.2) | 1.42 (0.2) | 1.58 (0.3) | 6.53$\times$10^-15^ |
| Amyloid PET, No. (%) | | |  |  |  |  |
|  | Negative | | 218 (49.3) | 61 (27.2) | 9 (15.3) | 6.59$\times$10^-10^ |
|  | Positive | | 190 (43.0) | 128 (57.1) | 46 (78.0) |  |
| MMSE, mean (SD) | | | 27.46 (2.5) | 25.93 (2.9) | 17.63 (6.8) | 9.18$\times$10^-79^ |
| SNSB: Language, mean (SD) | | | 0.18 (0.02) | -0.12 (0.03) | -0.48 (0.06) | 3.19$\times$10^-31^ |
| SNSB: Visuspatial, mean (SD) | | | 0.47 (0.03) | 0.10 (0.04) | -0.71 (0.09) | 5.63$\times$10^-37^ |
| SNSB: Memory, mean (SD) | | | 0.21 (0.03) | -0.51 (0.04) | -1.49 (0.08) | 3.04$\times$10^-88^ |
| SNSB: Executive function, mean (SD) | | | 0.20 (0.03) | -0.28 (0.04) | -1.00 (0.07) | 5.76$\times$10^-58^ |
| Left hippocampal volume, mean (SD) | | | 3726.23 (400.6) | 3493.55 (461.7) | 2989.89 (387) | 8.72$\times$10^-35^ |
| Right hippocampal volume, mean (SD) | | | 3916.7 (431.6) | 3699.37 (509.6) | 3138.43 (447.4) | 9.13$\times$10^-32^ |
| Left entorhinal thickness, mean (SD) | | | 3.29 (0.3) | 3.1 (0.4) | 2.7 (0.4) | 3.06$\times$10^-24^ |
| Right entorhinal thickness, mean (SD) | | | 3.44 (0.3) | 3.26 (0.4) | 2.79 (0.5) | 1.56$\times$10^-25^ |
| Medication usage^#^, No. (%) | | | 206 (46.6) | 96 (42.9) | 14 (23.7) | 0.004 |
| pTau217, mean (SD), pg/ml | | | 0.53 (0.4) | 0.68 (0.5) | 1.05 (0.5) | 4.81$\times$10^-17^ |
| GFAP, mean (SD), pg/ml | | | 242.79 (114.5) | 272.27 (124.3) | 366.99 (175.4) | 1.88$\times$10^-10^ |
| NfL, mean (SD), pg/ml | | | 23.11 (10.0) | 24.64 (9.7) | 28.71 (11.1) | 0.001 |

Abbreviations: CU, cognitive unimpaired; MCI, mild cognitive impairment; AD, Alzheimer’s disease; SD, standard deviation; MMSE, mini-mental state examination; SUVr, standard uptake value ratio. ^#^Medications related to hyperlipidemia or diabetes. Bold p-values indicate Bonferroni corrected p<0.005.

**Supplementary Table 2. Association with endophenotypes for AD status and Amyloid positivity as binary outcomes**

| **Endophenotype** | **CU vs. AD** | |  | **Amyloid positive vs. negative** | |
| --- | --- | --- | --- | --- | --- |
|  | **B (SE)** | **P** |  | **B (SE)** | **P** |
| **Cognitive Domains:** |  |  |  |  |  |
| Attention | -0.501 (0.07) | **3.51E-10** |  | -0.087 (0.03) | 0.015 |
| Language | -2.505 (0.35) | **3.14E-12** |  | -0.511 (0.18) | 0.006 |
| Visuospatial | -2.05 (0.26) | **1.47E-14** |  | -0.299 (0.11) | 0.010 |
| Memory | -4.942 (0.67) | **1.97E-13** |  | -0.657 (0.1) | **1.60E-9** |
| Executive function | -2.792 (0.33) | **2.64E-17** |  | -0.579 (0.11) | **1.09E-6** |
| **MRI Scans:** |  |  |  |  |  |
| Left hippocampal volume | -0.004 (0.001) | **1.39E-17** |  | 0.002 (0.001) | 0.008 |
| Right hippocampal volume | -0.003 (0.001) | **5.88E-17** |  | 0.003 (0.011) | 0.007 |
| Left entorhinal thickness | -3.183 (0.38) | **2.16E-16** |  | -0.376 (0.18) | 0.040 |
| Right entorhinal thickness | -2.958 (0.35) | **3.54E-17** |  | -0.359 (0.17) | 0.037 |

Abbreviations: CU, cognitive unimpaired; AD, Alzheimer’s disease; B, beta estimate; SE, standard error; P, p value. Bold p-values indicate Bonferroni corrected p<0.005.

**Supplementary Table 3. Association for pTau217 and AD related endophenotypes in all, amyloid positive, and negative groups**

| **Endophenotype** | **All** | | |  | **Amyloid positive** | | |  | **Amyloid negative** | | |
| --- | --- | --- | --- | --- | --- | --- | --- | --- | --- | --- | --- |
|  | **B** | **SE** | **P** |  | **B** | **SE** | **P** |  | **B** | **SE** | **P** |
| SNSB: Attention | -0.028 | 0.007 | **1.00E-4** |  | -0.030 | 0.010 | **0.004** |  | -0.006 | 0.004 | 0.123 |
| SNSB: Language | -0.179 | 0.034 | **1.43E-7** |  | -0.178 | 0.044 | **5.63E-5** |  | -0.041 | 0.022 | 0.066 |
| SNSB: Visuospatial | -0.117 | 0.021 | **3.06E-8** |  | -0.108 | 0.028 | **1.54E-4** |  | -0.029 | 0.014 | 0.039 |
| SNSB: Memory | -0.170 | 0.019 | **2.72E-18** |  | -0.152 | 0.027 | **2.50E-8** |  | -0.032 | 0.013 | 0.012 |
| SNSB: Executive function | -0.158 | 0.021 | **1.22E-13** |  | -0.145 | 0.029 | **6.64E-7** |  | -0.041 | 0.015 | 0.005 |
| Left hippocampal volume | -2.13E-4 | 3.44E-5 | **9.97E-10** |  | -2.46E-4 | 4.63E-5 | **1.95E-7** |  | -6.07E-5 | 2.12E-5 | 0.005 |
| Right hippocampal volume | -2.08E-4 | 3.15E-5 | **8.53E-11** |  | -2.40E-4 | 4.16E-5 | **1.83E-8** |  | -4.10E-5 | 1.99E-5 | 0.041 |
| Left entorhinal thickness | -0.183 | 0.037 | **7.53E-7** |  | -0.226 | 0.050 | **9.37E-6** |  | -0.049 | 0.022 | 0.025 |
| Right entorhinal thickness | -0.171 | 0.035 | **1.24E-6** |  | -0.217 | 0.049 | **1.05E-5** |  | -0.024 | 0.020 | 0.244 |

Abbreviations: B, beta estimate; SE, standard error; P, p value. Bold p-values indicate Bonferroni corrected p<0.005.

**Supplementary Table 4. Association for GFAP and AD related endophenotypes in all, amyloid positive, and negative groups**

| **Endophenotype** | **All** | | |  | **Amyloid positive** | | |  | **Amyloid negative** | | |
| --- | --- | --- | --- | --- | --- | --- | --- | --- | --- | --- | --- |
|  | **B** | **SE** | **P** |  | **B** | **SE** | **P** |  | **B** | **SE** | **P** |
| SNSB: Attention | -5.645 | 2.141 | 0.009 |  | -6.703 | 3.158 | 0.035 |  | -1.723 | 2.198 | 0.434 |
| SNSB: Language | -37.507 | 10.285 | **2.86E-4** |  | -45.048 | 14.154 | **0.002** |  | -8.780 | 11.550 | 0.448 |
| SNSB: Visuospatial | -29.593 | 6.432 | **5.01E-6** |  | -38.799 | 9.258 | **3.56E-5** |  | -2.785 | 7.329 | 0.704 |
| SNSB: Memory | -41.543 | 5.776 | **1.69E-12** |  | -48.307 | 8.341 | **1.60E-8** |  | -6.119 | 6.665 | 0.359 |
| SNSB: Executive function | -35.781 | 6.310 | **2.11E-8** |  | -37.160 | 9.031 | **4.88E-5** |  | -10.687 | 7.555 | 0.158 |
| Left hippocampal volume | -0.059 | 0.010 | **1.82E-8** |  | -0.074 | 0.015 | **8.70E-7** |  | -0.026 | 0.011 | 0.019 |
| Right hippocampal volume | -0.060 | 0.009 | **4.64E-10** |  | -0.076 | 0.013 | **1.53E-8** |  | -0.019 | 0.011 | 0.077 |
| Left entorhinal thickness | -55.303 | 11.023 | **6.70E-7** |  | -75.188 | 15.983 | **3.72E-6** |  | -22.076 | 11.361 | 0.053 |
| Right entorhinal thickness | -48.484 | 10.445 | **4.15E-6** |  | -66.116 | 15.403 | **2.31E-5** |  | -19.763 | 10.416 | 0.059 |

Abbreviations: B, beta estimate; SE, standard error; P, p value. Bold p-values indicate Bonferroni corrected p<0.005.

**Supplementary Table 5. Association for NfL and AD related endophenotypes in all, amyloid-positive, and amyloid-negative subjects**

| **Endophenotype** | **All** | | |  | **Amyloid positive** | | |  | **Amyloid negative** | | |
| --- | --- | --- | --- | --- | --- | --- | --- | --- | --- | --- | --- |
|  | **B** | **SE** | **P** |  | **B** | **SE** | **P** |  | **B** | **SE** | **P** |
| SNSB: Attention | -0.219 | 0.178 | 0.218 |  | -0.267 | 0.245 | 0.278 |  | 0.040 | 0.266 | 0.880 |
| SNSB: Language | -2.703 | 0.840 | 0.001 |  | -3.583 | 1.053 | **7.45E-4** |  | -0.207 | 1.426 | 0.885 |
| SNSB: Visuospatial | -1.784 | 0.527 | **7.51E-4** |  | -1.688 | 0.692 | 0.015 |  | -0.462 | 0.921 | 0.617 |
| SNSB: Memory | -2.294 | 0.487 | **3.00E-6** |  | -2.285 | 0.659 | **5.90E-4** |  | -0.960 | 0.821 | 0.243 |
| SNSB: Executive function | -2.661 | 0.526 | **5.36E-7** |  | -2.055 | 0.693 | **0.003** |  | -2.352 | 0.919 | 0.011 |
| Left hippocampal volume | -0.003 | 0.001 | **3.34E-4** |  | -0.003 | 0.001 | 0.015 |  | -0.003 | 0.001 | 0.037 |
| Right hippocampal volume | -0.003 | 0.001 | **8.49E-5** |  | -0.003 | 0.001 | **0.001** |  | -0.003 | 0.001 | 0.048 |
| Left entorhinal thickness | -3.703 | 0.906 | **4.92E-5** |  | -3.746 | 1.244 | 0.003 |  | -2.599 | 1.383 | 0.061 |
| Right entorhinal thickness | -3.035 | 0.863 | **4.67E-4** |  | -4.577 | 1.185 | **1.35E-4** |  | -0.861 | 1.275 | 0.500 |

Abbreviations: B, beta estimate; SE, standard error; P, p value. Bold p-values indicate Bonferroni corrected p<0.005.

**Supplementary Table 6. Prediction of AD status using plasma biomarkers in all subjects**

|  | **CU vs. AD** | |  | **CU vs. MCI** | |  | **MCI vs. AD** | |
| --- | --- | --- | --- | --- | --- | --- | --- | --- |
|  | **AUC (95% CI)** | **P** |  | **AUC (95% CI)** | **P** |  | **AUC (95% CI)** | **P** |
| Demographics | 0.613 (0.52-0.70) | 1.51E-02 |  | 0.531 (0.48-0.58) | 2.37E-01 |  | 0.623 (0.53-0.72) | 1.24E-02 |
| +pTau217 | 0.790 (0.72-0.86) | 4.54E-10 |  | 0.595 (0.54-0.65) | 3.10E-04 |  | 0.713 (0.62-0.81) | 1.53E-05 |
| +GFAP | 0.703 (0.61-0.79) | 1.22E-05 |  | 0.578 (0.53-0.63) | 3.36E-03 |  | 0.690 (0.59-0.79) | 1.10E-04 |
| +NfL | 0.673 (0.59-0.76) | 2.00E-04 |  | 0.559 (0.51-0.61) | 2.60E-02 |  | 0.652 (0.56-0.75) | 2.07E-03 |
| +pTau217+GFAP | 0.777 (0.70-0.85) | 2.39E-09 |  | 0.598 (0.55-0.65) | 2.10E-04 |  | 0.707 (0.61-0.80) | 2.68E-05 |
| +pTau217+NfL | 0.798 (0.73-0.86) | 1.33E-10 |  | 0.600 (0.55-0.65) | 1.50E-04 |  | 0.723 (0.64-0.81) | 6.28E-06 |
| +GFAP+NfL | 0.716 (0.63-0.80) | 3.45E-06 |  | 0.580 (0.53-0.63) | 2.56E-03 |  | 0.698 (0.60-0.79) | 5.94E-05 |
| +pTau217+GFAP+NfL | 0.792 (0.72-0.86) | 3.12E-10 |  | 0.601 (0.55-0.65) | 1.30E-04 |  | 0.715 (0.62-0.81) | 1.30E-05 |

Abbreviations: CU, cognitive unimpaired; MCI, mild cognitive impairment; AD, Alzheimer’s disease; AUC, area under curve; CI, confidence interval; P, p value; Demo, demographics including age, sex and education.

**Supplementary Table 7. Association of blood biochemical markers for AD status and amyloid positivity in all subjects**

| **Blood biochemical marker** | **Diagnosis** | | | |  | **CU vs. AD** | |  | **Amyloid positive vs. negative** | |
| --- | --- | --- | --- | --- | --- | --- | --- | --- | --- | --- |
|  | **CU** | **MCI** | **AD** | **P** |  | **B (SE)** | **P** |  | **B (SE)** | **P** |
| Thrombocyte, mean (SE) | 210.43 (53.9) | 225.52 (55.9) | 225.81 (67.7) | 0.009 |  | 0.003 (0.003) | 0.261 |  | 0.0004 (0.002) | 0.793 |
| Hemoglobin, mean (SE) | 13.5 (1.2) | 13.16 (1.3) | 13.42 (1.3) | 0.026 |  | 0.017 (0.14) | 0.906 |  | -0.114 (0.08) | 0.892 |
| Hematocrit, mean (SE) | 40.74 (3.5) | 39.94 (3.6) | 40.56 (4.1) | 0.077 |  | 0.006 (0.05) | 0.893 |  | -0.022 (0.03) | 0.413 |
| Cholesterol, mean (SE) | 165.86 (37.1) | 167.93 (37.6) | 191.65 (42.7) | **5.04E-5** |  | 0.015 (0.01) | **0.002** |  | -0.001 (0.003) | 0.999 |
| Triglyceride, mean (SE) | 104.7 (47.9) | 109.78 (51.3) | 130.77 (49.3) | 0.003 |  | 0.008 (0.003) | 0.004 |  | -0.001 (0.002) | 0.473 |
| AST, mean (SE) | 27.24 (9.6) | 27.2 (12.6) | 27.89 (16.1) | 0.924 |  | 0.008 (0.01) | 0.524 |  | 0.001 (0.008) | 0.889 |
| ALT, mean (SE) | 19.78 (10.5) | 20.15 (12.5) | 18.83 (10) | 0.770 |  | 0.001 (0.02) | 0.928 |  | -0.003 (0.008) | 0.684 |
| BUN, mean (SE) | 17.87 (14.2) | 17.6 (6.1) | 16.73 (6.2) | 0.891 |  | -0.040 (0.05) | 0.375 |  | 0.014 (0.02) | 0.446 |
| Creatinine, mean (SE) | 0.81 (0.2) | 0.84 (0.2) | 0.83 (0.2) | 0.428 |  | 1.601 (1.1) | 0.157 |  | 0.639 (0.56) | 0.258 |
| HbA1c, mean (SE) | 5.94 (0.6) | 6.12 (0.8) | 6.11 (1) | 0.029 |  | 0.602 (0.2) | 0.003 |  | 0.115 (0.13) | 0.376 |
| LDL, mean (SE) | 97.1 (33) | 99.57 (35.8) | 121.36 (40.7) | **2.80E-5** |  | 0.015 (0.01) | **0.002** |  | -0.0005 (0.003) | 0.873 |
| HDL, mean (SE) | 54.2 (14.6) | 52.4 (14.9) | 53.35 (15.9) | 0.458 |  | -0.0003 (0.01) | 0.982 |  | 0.00006 (0.006) | 0.992 |
| Glucose, mean (SE) | 76.73 (20.2) | 79.99 (23.1) | 89.56 (33.2) | **0.001** |  | 0.033 (2.9) | **3.16E-6** |  | -0.001 (0.004) | 0.839 |

Abbreviations: AST, aspartate aminotransferase; ALT, alanine transaminase; BUN, blood urea nitrogen; LDL, low-density lipoprotein cholesterol; HDL, high-density lipoprotein cholesterol; CU, cognitive unimpaired; MCI, mild cognitive impairment; AD, Alzheimer’s disease; B, beta estimate; SE, standard error; P, p value. Statistically significant difference was shown in bold (p<0.003).

**Supplementary Table 8. Predictive performance in ROC curves of AD status using blood biochemical markers in all subjects**

| **Model** | **CU vs. AD** | |  | **CU vs. MCI** | |  | **MCI vs. AD** | |
| --- | --- | --- | --- | --- | --- | --- | --- | --- |
|  | **AUC (95% CI)** | **P** |  | **AUC (95% CI)** | **P** |  | **AUC (95% CI)** | **P** |
| Age | 0.591 (0.486-0.696) | 0.089 |  | 0.529 (0.471-0.586) | 0.320 |  | 0.443 (0.338-0.549) | 0.292 |
| Sex | 0.524 (0.434-0.615) | 0.591 |  | 0.509 (0.453-0.564) | 0.744 |  | 0.534 (0.436-0.632) | 0.494 |
| Education | 0.591 (0.498-0.683) | 0.053 |  | 0.521 (0.467-0.575) | 0.429 |  | 0.616 (0.515-0.718) | 0.025 |
| Demo (age+sex+education) | 0.587 (0.495-0.678) | 0.061 |  | 0.535 (0.481-0.590) | 0.195 |  | 0.607 (0.516-0.699) | 0.022 |
| Cholesterol | 0.662 (0.572-0.752) | 4.20E-04 |  | 0.522 (0.466-0.578) | 0.444 |  | 0.640 (0.543-0.736) | 0.005 |
| Glucose | 0.615 (0.520-0.710) | 0.018 |  | 0.550 (0.493-0.606) | 0.086 |  | 0.569 (0.467-0.670) | 0.184 |
| Demo+Med | 0.634 (0.552-0.716) | 0.001 |  | 0.552 (0.497-0.606) | 0.059 |  | 0.607 (0.516-0.699) | 0.047 |
| +Cholesterol | 0.682 (0.597-0.767) | 2.62E-05 |  | 0.555 (0.499-0.611) | 0.054 |  | 0.669 (0.572-0.766) | 6.43E-04 |
| +Glucose | 0.736 (0.662-0.809) | 2.96E-10 |  | 0.588 (0.532-0.644) | 0.002 |  | 0.680 (0.590-0.770) | 8.58E-05 |
| +Cholesterol+Glucose | 0.765 (0.695-0.835) | 1.31E-13 |  | 0.588 (0.532-0.643) | 0.002 |  | 0.712 (0.623-0.802) | 3.53E-06 |

Abbreviations: CU, cognitive unimpaired; MCI, mild cognitive impairment; AD, Alzheimer’s disease; AUC, area under curve; CI, confidence interval; P, p value; Demo, demographics including age, sex, and education; Med, medications related to hyperlipidemia or diabetes.

**Supplementary Table 9. Predictive performance in ROC curves of AD status using pTau217 and blood biochemical markers in amyloid positive subgroup**

| **Model** | **CU+ vs. AD+** | |  | **CU+ vs. MCI+** | |  | **MCI+ vs. AD+** | |
| --- | --- | --- | --- | --- | --- | --- | --- | --- |
|  | **AUC (95% CI)** | **P** |  | **AUC (95% CI)** | **P** |  | **AUC (95% CI)** | **P** |
| Age | 0.585 (0.458-0.713) | 1.85E-01 |  | 0.536 (0.459-0.613) | 3.55E-01 |  | 0.460 (0.335-0.585) | 5.38E-01 |
| Sex | 0.537 (0.426-0.647) | 5.09E-01 |  | 0.532 (0.457-0.608) | 3.96E-01 |  | 0.495 (0.378-0.612) | 9.40E-01 |
| Education | 0.565 (0.451-0.679) | 2.63E-01 |  | 0.557 (0.483-0.631) | 1.26E-01 |  | 0.621 (0.498-0.744) | 5.40E-02 |
| Demo (age+sex+education) | 0.566 (0.449-0.683) | 2.66E-01 |  | 0.590 (0.516-0.663) | 1.60E-02 |  | 0.646 (0.529-0.763) | 1.40E-02 |
| +pTau217 | 0.755 (0.669-0.841) | 5.32E-09 |  | 0.585 (0.510-0.660) | 2.60E-02 |  | 0.725 (0.620-0.830) | 2.51E-05 |
| +GFAP | 0.682 (0.574-0.791) | 1.00E-03 |  | 0.594 (0.519-0.668) | 1.30E-02 |  | 0.698 (0.584-0.812) | 1.00E-03 |
| +NfL | 0.681 (0.580-0.781) | 4.10E-04 |  | 0.606 (0.533-0.680) | 5.00E-03 |  | 0.656 (0.541-0.770) | 7.00E-03 |
| Demo+Med | 0.605 (0.500-0.710) | 4.80E-02 |  | 0.578 (0.504-0.652) | 3.80E-02 |  | 0.615 (0.508-0.723) | 3.40E-02 |
| +pTau217 | 0.777 (0.694-0.860) | 5.74E-11 |  | 0.575 (0.500-0.651) | 4.95E-02 |  | 0.737 (0.639-0.835) | 2.17E-06 |
| +GFAP | 0.702 (0.603-0.801) | 5.93E-05 |  | 0.581 (0.507-0.656) | 3.19E-02 |  | 0.707 (0.599-0.815) | 1.59E-04 |
| +NfL | 0.689 (0.594-0.784) | 9.59E-05 |  | 0.597 (0.523-0.671) | 1.01E-02 |  | 0.648 (0.543-0.754) | 5.52E-03 |
| +Cholesterol | 0.654 (0.540-0.768) | 0.008 |  | 0.602 (0.529-0.676) | 0.006 |  | 0.649 (0.534-0.764) | 0.011 |
| +Glucose | 0.783 (0.698-0.868) | 7.39E-11 |  | 0.661 (0.590-0.733) | 8.30E-06 |  | 0.696 (0.596-0.797) | 1.21E-04 |
| +Glucose+pTau217 | 0.861 (0.796-0.926) | 1.00E-27 |  | 0.674 (0.603-0.745) | 1.34E-06 |  | 0.784 (0.697-0.872) | 1.68E-10 |
| +Glucose+GFAP | 0.812 (0.733-0.891) | 9.55E-15 |  | 0.658 (0.587-0.729) | 1.20E-05 |  | 0.744 (0.644-0.844) | 1.65E-06 |
| +Glucose+NfL | 0.798 (0.716-0.880) | 1.13E-12 |  | 0.663 (0.593-0.732) | 4.32E-06 |  | 0.710 (0.610-0.810) | 3.51E-05 |
| +Glucose+pTau217+GFAP | 0.859 (0.793-0.925) | 1.50E-26 |  | 0.668 (0.597-0.739) | 3.12E-06 |  | 0.773 (0.683-0.864) | 3.10E-09 |
| +Glucose+GFAP+NfL | 0.819 (0.740-0.897) | 1.55E-15 |  | 0.661 (0.592-0.731) | 5.21E-06 |  | 0.747 (0.647-0.847) | 1.24E-06 |
| +Glucose+Cholesterol | 0.802 (0.722-0.883) | 1.98E-13 |  | 0.663 (0.592-0.734) | 6.90E-06 |  | 0.710 (0.602-0.817) | 1.31E-04 |
| +Glucose+Cholesterol+GFAP | 0.828 (0.749-0.908) | 4.44E-16 |  | 0.658 (0.587-0.729) | 1.29E-05 |  | 0.771 (0.666-0.877) | 4.57E-07 |
| +Glucose+Cholesterol+NfL | 0.815 (0.736-0.894) | 5.77E-15 |  | 0.670 (0.601-0.739) | 1.42E-06 |  | 0.719 (0.608-0.830) | 1.05E-04 |
| +pTau217+NfL | 0.785 (0.710-0.860) | 1.03E-13 |  | 0.590 (0.516-0.665) | 1.70E-02 |  | 0.736 (0.638-0.835) | 2.31E-06 |
| +pTau217+GFAP | 0.777 (0.697-0.857) | 1.35E-11 |  | 0.576 (0.500-0.652) | 4.80E-02 |  | 0.739 (0.641-0.838) | 1.87E-06 |
| +pTau217+GFAP+NfL | 0.782 (0.706-0.858) | 3.04E-13 |  | 0.586 (0.511-0.660) | 2.43E-02 |  | 0.739 (0.639-0.839) | 2.54E-06 |
| +pTau217+Glucose+Cholesterol | 0.867 (0.803-0.932) | 2.00E-28 |  | 0.675 (0.605-0.746) | 1.11E-06 |  | 0.793 (0.701-0.884) | 4.18E-10 |
| +pTau217+Glucose+Cholesterol+NfL | 0.868 (0.805-0.932) | 1.00E-28 |  | 0.672 (0.602-0.741) | 1.24E-06 |  | 0.788 (0.695-0.882) | 1.38E-09 |
| +pTau217+Glucose+Cholesterol+GFAP | 0.869 (0.806-0.932) | 8.00E-29 |  | 0.669 (0.598-0.740) | 2.67E-06 |  | 0.792 (0.697-0.887) | 1.48E-09 |
| +pTau217+Glucose+Cholesterol+GFAP+NfL | 0.868 (0.805-0.932) | 6.00E-30 |  | 0.672 (0.602-0.741) | 1.31E-06 |  | 0.789 (0.693-0.885) | 3.93E-09 |

Abbreviations: AUC, area under curve; CI, confidence interval; P, p value; Demo, demographics including age, sex, and education; Med, medications related to hyperlipidemia or diabetes. Amyloid-positive individuals (+) from cognitive unimpaired (CU), mild cognitive impairment (MCI), and Alzheimer’s disease (AD).

**Supplementary Table 10. Conversion to AD dementia during follow-up**

| **Baseline diagnosis** | **N** | **Converted to AD, n (%)** | **Years of interval, mean (SD)** |
| --- | --- | --- | --- |
| CU- | 145 | 0 (0) | 5.68 (2.99) |
| CU+ | 169 | 4 (2.37) | 5.56 (2.63) |
| MCI- | 12 | 1 (8.33) | 4.48 (2.50) |
| MCI+ | 74 | 14 (18.92) | 4.46 (2.55) |

Abbreviations: N, number of samples; SD, standard deviation; CU, cognitive unimpaired; MCI, mild cognitive impairment; AD, Alzheimer’s disease; +, amyloid positive; −, amyloid negative.

**Supplementary Table 11. AD conversion analysis using pTau217 and blood biochemical markers in amyloid positive subgroup**

| **Model** | **AUC (95% CI)** | **P** |
| --- | --- | --- |
| Age | 0.678 (0.521-0.835) | 0.026 |
| Sex | 0.622 (0.480-0.765) | 0.091 |
| Education | 0.602 (0.437-0.766) | 0.225 |
| Demo (age+sex+education) | 0.719 (0.564-0.875) | 0.006 |
| +pTau217 | 0.734 (0.594-0.874) | 0.001 |
| +GFAP | 0.763 (0.625-0.901) | 1.88E-04 |
| +NfL | 0.730 (0.579-0.881) | 0.003 |
| Demo+Med | 0.732 (0.580-0.885) | 0.003 |
| +pTau217 | 0.747 (0.612-0.882) | 3.13E-04 |
| +NfL | 0.738 (0.585-0.891) | 2.26E-03 |
| +GFAP | 0.767 (0.633-0.901) | 8.79E-05 |
| +Cholesterol | 0.732 (0.578-0.886) | 3.06E-03 |
| +Glucose | 0.840 (0.754-0.925) | 6.88E-15 |
| +Glucose+pTau217 | 0.831 (0.741-0.922) | 7.79E-13 |
| +Glucose+GFAP | 0.867 (0.789-0.944) | 2.80E-20 |
| +Glucose+NfL | 0.845 (0.761-0.930) | 8.88E-16 |
| +Glucose+pTau217+GFAP | 0.867 (0.790-0.943) | 5.00E-20 |
| +Glucose+GFAP+NfL | 0.870 (0.791-0.949) | 1.00E-20 |
| +Glucose+Cholesterol | 0.826 (0.733-0.919) | 6.75E-12 |
| +Glucose+Cholesterol+GFAP | 0.868 (0.792-0.944) | 3.50E-20 |
| +Glucose+Cholesterol+NfL | 0.846 (0.764-0.929) | 1.20E-16 |
| +pTau217+GFAP+NfL | 0.765 (0.632-0.898) | 9.26E-05 |
| +pTau217+Cholesterol+Glucose | 0.828 (0.736-0.920) | 2.08E-12 |
| +pTau217+NfL | 0.742 (0.594-0.890) | 0.001 |
| +pTau217+GFAP | 0.764 (0.630-0.899) | 1.14E-04 |
| +pTau217+Glucose+Cholesterol+NfL | 0.843 (0.758-0.928) | 1.78E-15 |
| +pTau217+Glucose+Cholesterol+GFAP | 0.867 (0.792-0.943) | 1.60E-21 |
| +pTau217+Glucose+Cholesterol+GFAP+NfL | 0.874 (0.798-0.950) | 4.00E-22 |

Abbreviations: AUC, area under curve; CI, confidence interval; P, p value; Demo, demographics including age, sex, and education; Med, medications related to hyperlipidemia or diabetes. Amyloid-positive individuals (+) from cognitive unimpaired (CU), mild cognitive impairment (MCI), and Alzheimer’s disease (AD). AD conversion status was defined as a binary outcome, where a value of 1 indicated conversion from CU+ or MCI+ to AD+, and 0 indicated that the CU+ or MCI+ diagnosis remained unchanged at the final examination.
